# State-dependent patterns and coupling between the vaccination schedule, population mobility and the COVID epidemic outline, in the US states

**DOI:** 10.1101/2021.07.18.21260708

**Authors:** Anca Rǎdulescu, Tatiana Alonso, Norman Reid, Johanna Sanchez

## Abstract

We study the evolution of the COVID-19 epidemic in the US, since January 2020 until May 2021. Our primary goal is to understand some of the complex coupled dynamics between factors that ultimately regulate the epidemic case load. As potentially crucial factors, we focus on population mobility and vaccination patters (both related to risk of contracting the SARS-Cov2 virus). These factors may in turn depend on demographic parameters (which are unrelated to the epidemic evolution), but also on the population response to the epidemic outbreak itself. In our work, we use correlation analyses, in conjunction with open source data from US states, to investigate the type and strength (1) of the relationships between demographic measures and epidemic, mobility and vaccination timelines during our established time window; (2) of the bidirectional coupling between these timelines.

We showed that the wide between-state differences in epidemic outcome correspond to between-state differences in demographic measures (such as density, income, political affiliation). As a potential underlying mechanism, we found that demographic measures are also predictive of the degree of coupling between epidemic timelines (on one hand) and vaccination and mobility timelines (on the other hand), coupling which can be broadly interpreted as the population’s behavioral response to the epidemic. In support of this idea, our analysis shows this response to be tightly correlated with epidemic outcome.

This suggests that a state’s demographic profile may be invaluable to generating predictions on the epidemic evolution in the respective state, and that this information may be used to understand the weaknesses of a state and how to compensate for them to improve epidemic outcome (e.g., via state centralized incentives, and customized mitigation strategies).

## 1 Introduction

The COVID-19 epidemic has remained a crucial public concern for longer than a year, affecting the lives, health and livelihood of millions of people all around the world. When discussing the dramatic development of the COVID-19 pandemic, a primary epidemiological concern is COVID-19 epidemic incidence, which is typically represented either as cumulative incidence (the total number of confirmed cases, useful when evaluating overall epidemic outcome), or as daily incidence (the number of daily confirmed cases, which is a useful measure when assessing change, or tracking the epidemic evolution over time). One crucial factor that impacts incidence is the spread rate (*R*_0_ number, which estimates in average the number of people that are infected by every person that is shedding SARS-CoV2 virus). In the case of SARS-Cov2, this number has been estimated up to *R*_0_ = 6 for its original form and variants [18]. Aside from this epidemiological profile of the virus itself, factors that are expected to dramatically impact incidence are exposure (which depends tightly on the degree of population mobility to various destinations [12, 19, 16], and on implementation and degree of social distancing measures [15, 14]) and, more recently, vaccinations. There factors are themselves influenced by demographic aspects [20], government regulations [16], the population’s behavioral response [13], as well as a variety of other (seemingly random) factors, including weather. In the diagram in Figure 1, we illustrate a few of the important interactions within the complex system in which COVID-19 incidence is only one of the variables.

**Figure 1:**
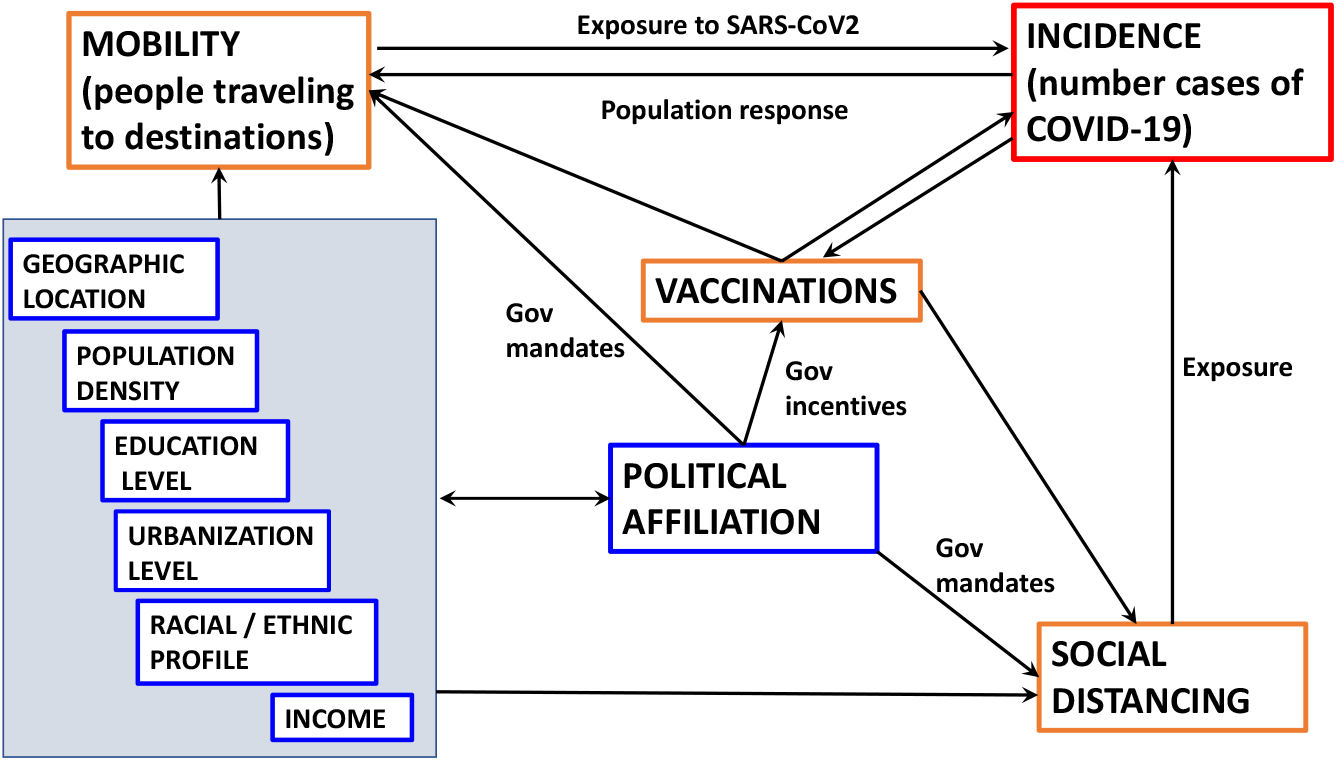
Subsystem of the self-interactive complex network that regulates epidemic incidence.

In our paper, we aim to understand some of these interactions (and potential driving mechanisms), based on state-level data on the evolution of the COVID-19 epidemic in the United States. In particular, we focus on the coupling of epidemics with mobility and vaccinations, as they evolved together in the US states. On one hand, population mobility [22, 17] and vaccinations are primary factors expected to impact epidemic outcome. On the other, they depend in turn on existing conditions in the corresponding state (which can partly quantified by measures such as population density, education level, political affiliation), and on the state of the epidemic itself (via the population behavioral response to risk of exposure [11]).

We investigate whether the epidemic, mobility and vaccination timelines and their interactions are related to the “demographic profile” of each state. Such relationships can be of utmost importance, since they relate quantitative aspects of phenomena which right now are very fluid and difficult to control, to more stable, slower changing, well-studied variables. Understanding the relationship between the two types of data could help generate better epidemiological predictions, as well as better assessments of government measure implementations (which are crucial for the control of this pandemic, but may be more generally applicable to other, future epidemic outbreaks).

The paper is organized as follows: in the Methods section, we introduce the source, type, pre-processing and limitations for each data set, and we present separately the trends in epidemics, mobility and vaccinations. In the Results section, we use correlation analyses to search for relationships across data sets, as potential markers for different coupling mechanisms that govern the evolution of the overall system. These relationships are interpreted and contextualized in the Discussion section.

## 2 Methods

We accessed epidemic, mobility and vaccination data from public reports and repositories, from the start of the pandemic until the end date of this study (May 22, 2021).

### 2.1 State demographic profiles

#### Census data

Since the 2020 census data is not yet available in the public domain, we based our computations on the 2019 US-wide estimate [9], which we found more appropriate to use than the previous (2010) US census. These estimates were used, whenever expressing a measure (e.g., vaccinations, or mortality) in terms of % of the state population.

**Population density** was computed for each state, using the 2019 census estimate, and the area of each state from [8].

#### Socioeconomic status

The median household income was obtained for each state from [10].

#### Political data

For each of the 51 states with existing procedures for US election, the political orientation was estimated from public data on voting in the 2020 presidential elections [1]. Since the blue (Democratic Party) and red (Republican party) options accounted for more than 95% in each one of the 51 states, we considered the % of blue voters to be an appropriate statistical variable to encompass the state’s political affiliation in 2020-2021 (i.e., for the duration of the pandemic to date).

#### Racial/ethnic profile

For each state [7], we obtained information regarding the % of the population which fell into one of the following categories: White, Black American, Hispanic, Asian, Native American (described as American Indian and Alaskan Native). These groups were not exhaustive, since our study did not include reports on Pacific Islanders, and people who identified as “multiple races.”

##### 2.1.1 Epidemic timelines

Epidemic data was accessed from New York Times reports [4], providing information on state-wide COVID-19 incidence and mortality, from January 12, 2020 until May 22, 2021 (501 days) [**?**]. Time series were created for each state, for four different measures associated with the COVID-19 epidemic: cumulative incidence (*CI*); percent cumulative incidence (%*CI*); daily incidence (*DI*); percent daily incidence (%*DI*).

Our first approach to the epidemic data included preprocessing (to eliminate artifacts and biases) and visualization (in order to guide our subsequent analysis efforts). We started by using the cumulative incidence *CI* (total number of confirmed COVID-19 cases in each state as of May 22, 2021), and percent cumulative incidence %*CI* (number of cases reported as a percent of the state population) to understand which sate had been most affected overall (ignoring the timing). We noticed that the impact on the states changes dramatically when considering these two different measures. Figure 2 illustrates as bar graphs the states in decreasing order of epidemic magnitude, when considering *CI* (panel A) and %*CI* (panel B). For example, California was the most affected state in terms of sheer numbers, but was not even among the top half states in terms of percent incidence (normalized by California’s very high population), whereas North Dakota, relatively low in *CI*, emerges as the most affected state in percent incidence (which accounts for North Dakota’s low population).

**Figure 2:**
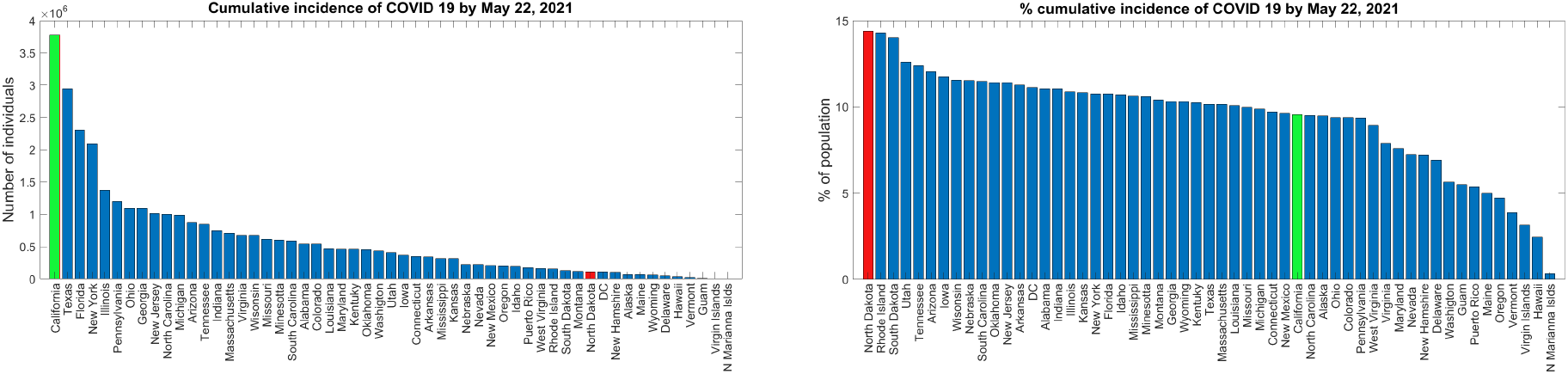
Bar graphs of US states in decreasing order of *CI* and %*CI*, respectively. *(as reported on May 22, 2021*. **A**. *When shown in decreasing order of CI, California shows the highest value (highlighted in green in both panels)*. **B**. *When shown in decreasing order of* %*CI, North Dakota shows the highest value (highlighted in red in both panels). All other states are shown in blue, and labeled*.

This encouraged the question of the relationship of %*CI* in each state with the state’s population density, as well as with other factors that may relate to population density (among which we considered political orientation, median household income (*MHI*) and racial/ethnic profile. To understand these relationships, we performed correlation analyses between %*CI* and these state demographic measures.

For our correlation analyses, we used a nonparametric test, the Spearman rank-order correlation (the Pearson correlation coefficient between rank variables). For two measures *X* and *Y*, this is defined as

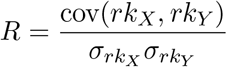

where *rk*_*X*_ and *rk*_*Y*_ are the rank variables corresponding to *X* and *Y*.

Spearman’s rank-order correlation assesses monotonic relationships (whether linear or not), with a positive value 0 < *R* ≤ 1 when *X* and *Y* tend to be simultaneously high, and a negative value −1 ≤ *R* < 0 if *Y* tends to be high when *X* is low. We used the Spearman correlation, because it is less sensitive than the Pearson correlation to strong outliers in the tails of both samples (since the outlier is limited to the value of its rank). In this study, we will report correlations to be statistically significant if the significance value *p* < 0.01.

We next included timing into the picture, and inspected the timelines of %*DI*(*k*) (where *k* is the day from January 12, 2000, until the end of our study) that lead to the %*CI* observed on May 22, 2021. The daily COVID-19 incidence timeline presented with fluctuations and irregularities which are not representative of the epidemic dynamics. These artifacts primarily reflect weekly seasonality in reporting (with an increase in case reports mid week, and a decrease during weekends) and abrupt adjustments to account for over- or under-reporting (visible as negative and respectively positive spikes in some of the state timelines). For the analyses where we only needed to tally up cumulative incidence over a time period, the precise occurrence of new cases is not expected to bring any significant change. However, for the sake of the analyses where the temporal evolution of the epidemic is in question, we smoothed out the daily incidence time series by calculating a 7-day moving window average. Throughout this study, only smoothed time series were used for illustration and analysis, hence we will henceforth use the notation *DI*(*k*) and %*DI*(*k*) for the smoothed time series, without danger of confusion. Figure 3 illustrates the percent daily incidence timecourse %*DI*(*k*) for the US as a whole; Figure 3b shows the %*DI*(*k*) time series for each individual state, with three states highlighted for example purposes.

**Figure 3:**
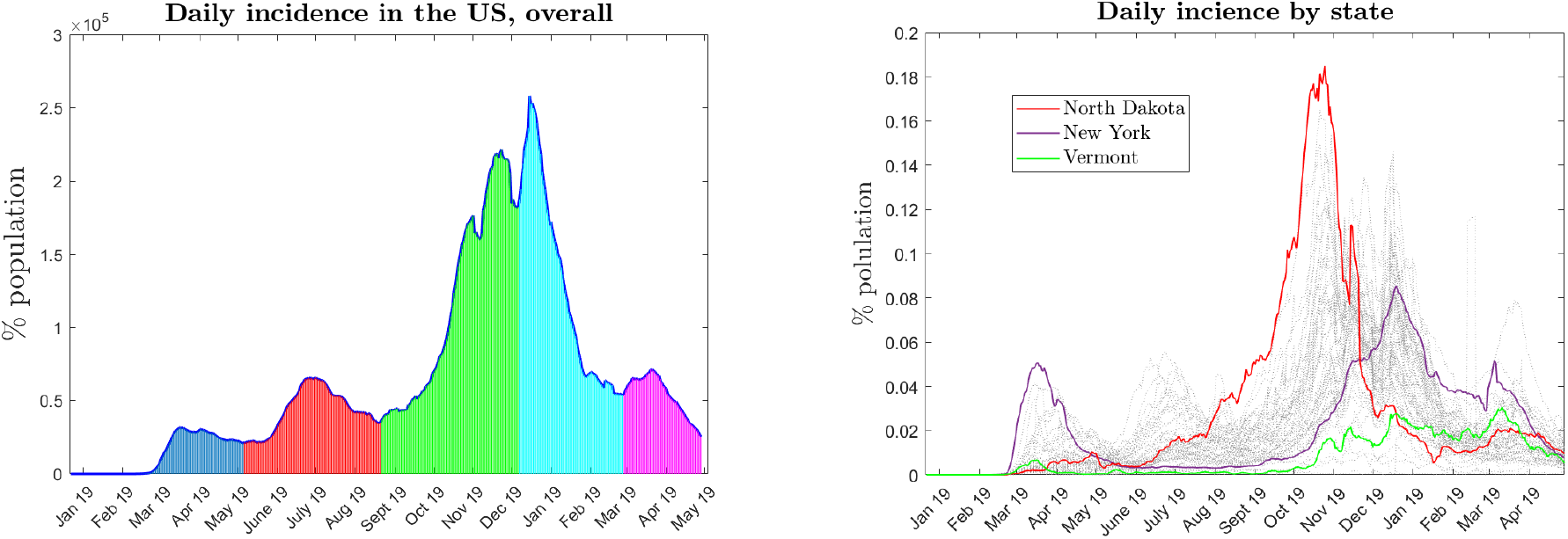
Percent daily incidence time courses in the United States, *from January 19, 2020, until May 22, 2021*. **A**. *Percent daily incidence* %*DI*(*k*), *for the US overall. To facilitate interpretation, the date (and not the k) is shown along the horizontal axis. The four waves (delimited as described in the text) are shaded in different colors*. **B**. %*DI time course is shown separately for each individual state, to illustrate the heterogeneity in between states in wave magnitude. Three states are highlighted in color, for illustration purposes: North Dakota (in red), which was the state with the highest* %*CI on May 22, 2021; New York (in purple), which was notoriously one of the states with the most prominent first wave; Vermont (in green), in which the infection was consistently low*.

Based on the shape of the US timeline in Figure 3a, but also on existing information on the evolution of COVID-19 cases in the US, and on the state-wide shapes shown in Figure 3b, we defined four major waves for the overall US epidemic timeline. Specificaly, we parsed these waves using the local minima of the graph in Figure 3a, as follows: first wave from January 8 until May 23, 2020 (136 days); second wave from May 24 until September 7, 2020 (107 days); third wave from September 8 until December 24, 2020 (108 days); fourth wave from December 25, 2020 until March 16, 2021 (82 days); fifth wave from March 17 until May 22, 2021 (61 days).

On the other hand, Figure 3b suggests that, while this wave partition was broadly consistent between states, states’ behavior was heterogeneous with respect to this broad structure, in the sense that different states experienced different waves, at very different magnitudes. Scientific discussions around the epidemic have questioned whether these waves had different underlying mechanisms, and to what extent these mechanisms could be explained by (or even predicted from) state existing demographic factors. To this aim, we computed the cumulative incidence for each wave from the smoothed *DI*(*k*) time series, and performed correlation analyses between this measure of the epidemic in each state, and the state’s demographic profile measures.

### 2.2 Vaccination timelines

From the pubic domain [5], we obtained for each state data on the population percent of fully vaccinated individuals (percent cumulative vaccinations, %*CV*, as of May 22, 2021), and time series for daily percents of vaccinated individuals (%*DV* (*k*), as reported for each day *k* from January 20, 2021, until the end of our study). The time series used from this open source had already been slightly preprocessed (e.g., smoothed).

Our first approach to these data was to inspect the efficiency of each state’s vaccination efforts, as expressed by %*CV*. Figure 4a shows that states exhibited a wide variation between 26-49.6% population fully vaccinated by May 22, 2021. We first aimed to check whether this efficiency correlated with the state’s demographic profile measures, which would help us understand which features within this profile could be predictive of high vaccination outcomes, and which states may need additional incentives to accomplish a more efficient scenario.

**Figure 4:**
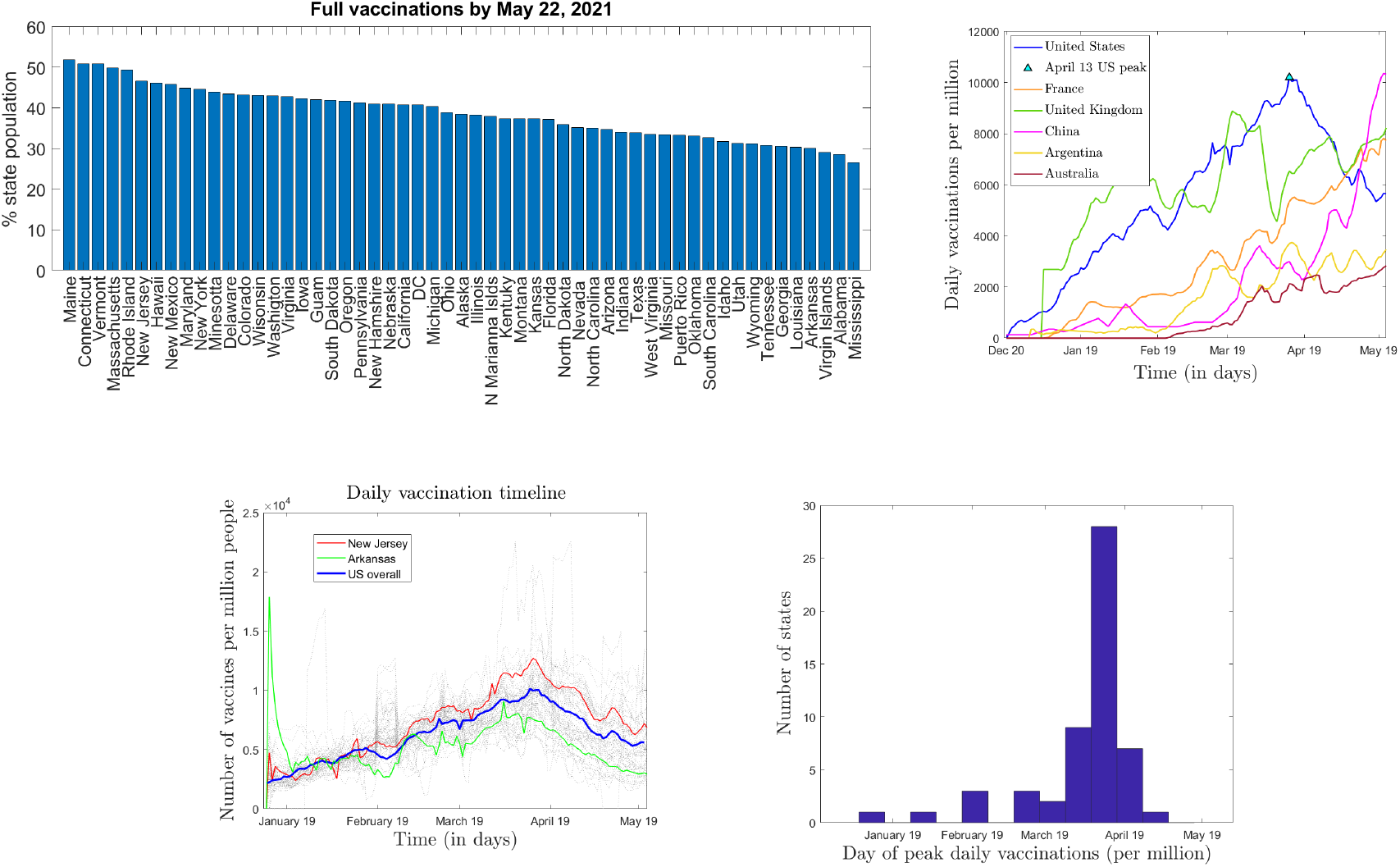
Presentation of vaccination data in the US states, *from December 20, 2020, until May 22, 2021*. **A**. *Bar graph of fully vaccinated population percent (*%*CV) on May 22, 2021, in the 55 US states for which data was available (shown in decreasing order)*. **B**. *Vaccination timeline (daily vaccinations per million, from December 20, 2020, until May 22, 2021) for the US (in blue) by comparison with a few other random countries, shown in other colors (as specified in the legend)*. **C**. *Vaccination timelines for all US states, from January 19 until May 22, 2021. Two states (New Jersey, in red, and Arkansas, in green) are shown as examples; the rest are all shown as dotted gray lines (highly cross-correlated). The timeline for the US overall is shown again in blue, as reference*. **D**. *Histogram of the day of occurrence of the peak vaccination rate, for each state. Most states peaked on or shortly after April 12; the few states that peaked early increased to another peak on or after April 12 (e*.*g*., *Arkansas)*.

Next, we considered the timing aspect, and analyzed the evolution of daily vaccinations that lead over time to the %*CV* observed by the end of the study, and shown in Figure 4a. We noticed a specific signature in the evolution of %*DV* (*k*) in the US, as illustrated in Figure 4b by comparison with a few other random countries. This signature consisted of an overall increase (via transient fluctuations) in percent daily vaccinations %*DV*, to a peak of 1% per day on April 12, 2021, followed by a consistent decay (also via transient fluctuations) until the end of our study on May 22, 2021. Since this shape and timing appear to be particular to the US, we first verified whether they are consistent among the 51 states for which vaccination data was provided. The illustration of the %*DV* (*k*) time series for all states, as shown in Figure 4c, suggests that this is the case. To quantify this idea, we also calculated mutual cross-correlations of the state-wide daily vaccination courses, and obtained a mean mutual cross-correlation value of 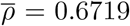, and a mean significance value 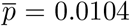. In addition, a histogram of the peak vaccination day (within the given time interval) for each state shows a very narrow temporal distribution of the time of the peaks. A few outliers of this distribution occurred early (as early as the second vaccination day, in the case of Arkansas, for example), but even these states peaked again around April 12, 2021.

This across-state consistency in the presence and timing of vaccination peak days suggests a triggering event. Indeed, on April 12, 2021, clinical side effects had been reported in conjunction with the Johnson and Johnson vaccine [6], subsequent to which the CDC and the FDA recommended pausing its administration, recommendation which was lifted on April 23 [3]. While is may be conceivable that a transient decay in vaccination rates may have occurred due to postponing the J&J count, the decreasing trend observed in data transcends this explanation, since it remains consistent even after the J&J vaccine had been reinstated. We would like to investigate the potential that the decrease in daily vaccinations may in fact be a behavioral response to the J&J incident. To follow up with this hypothesis, we investigated whether the extent to which vaccinations dropped in each state after achieving their peak (Δ%*DV*) was based on the state demographic profile. To this aim, we computed Spearman rank correlations between Δ%*DV* and the state demographic measures.

Finally, we wanted to study whether states with higher vaccination rates were more efficient at controlling the epidemic spread, once vaccinations started. To this aim, we computed correlations between the vaccination measures %*CV* and Δ%*DV*, and the epidemic measure %*CI*.

### 2.3 Traffic timelines

Time series describing traffic activity between January 13 and May 22, 2021 were made available in the public domain by Apple [2], based upon their daily numbers of hits for driving / walking / public transit directions requests. The accessible data reports the number of hits originating in each individual county as a percent of the baseline for that county. This baseline was defined by the county-wide number of hits on January 13, 2020, but was not explicitly provided. While the county-specific baseline makes magnitude comparisons meaningless between different counties, one can still compare the trends in the time series (e.g., one county had a steeper drop than another). Since the daily time series showed pronounced weekly oscillations, we obtained (as in the case of infection rates) smoothed time series, by using a seven day moving window average. One additional problem, which we will discuss later, is the absence of control time series. Since Apple did not make available any data from previous years, it is impossible to interpret the effect of seasonality or annual events.

Figure 5 illustrates side by side the smoothed driving timeline and the smoothed walking timeline for all states, between January 19, 2020 and May 22, 2021. After a significant drop initiated at the start of March 2020 (in conjunction with the first COVID-19 travel regulations and lockdowns), states began their reopening stages, and the timelines exhibit wide differences and fluctuations. A few states are highlighted in the two panels, to illustrate differences between states, as well as between different types of mobility for the same state.

**Figure 5:**
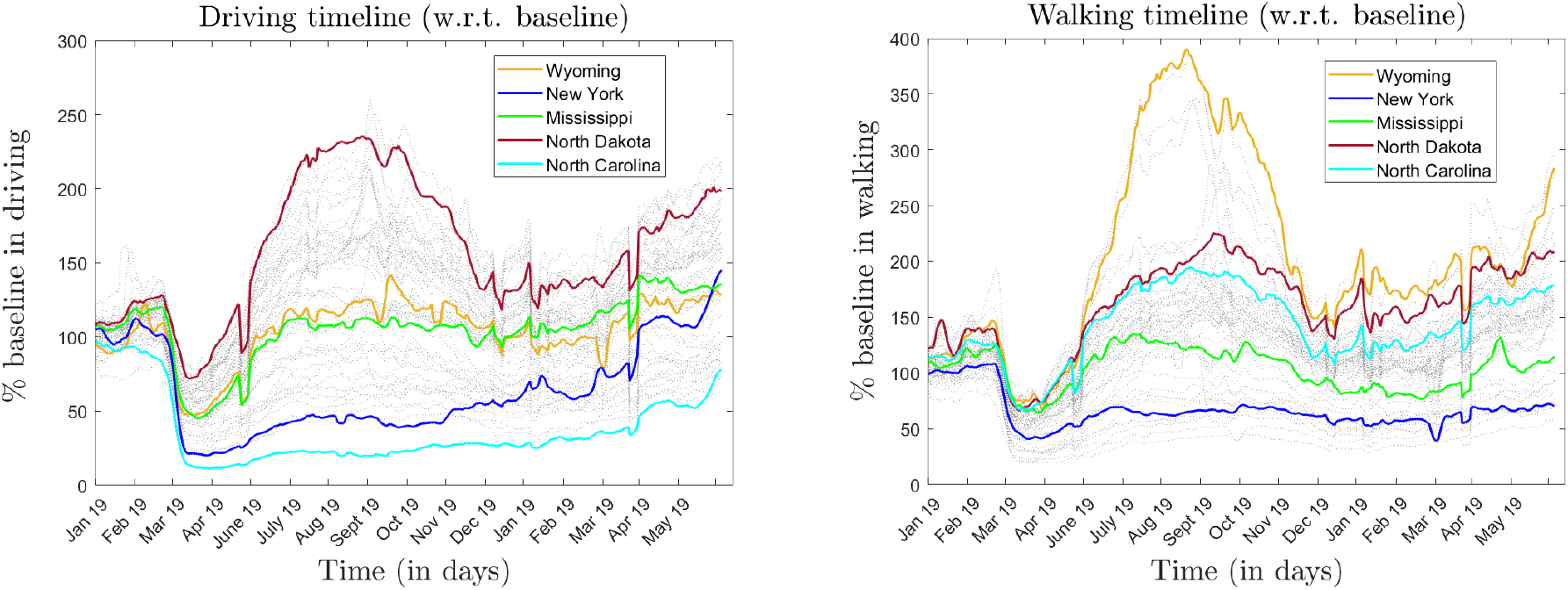
Smoothed traffic timelines for all states, *as provided by Apple (smoothed with a seven day moving window average). A few states are highlighted New York (blue); North Dakota (red); Mississippi (green); Missouri (pink); Arkansas (yellow); Wyoming (brown); Wisconsin (orange); North Carolina (cyan)*.

One interesting line of investigation would be to try to understand the factors and mechanisms behind these differences. Together with weather seasonality and weather fluctuations, one obvious influence to consider is that of the epidemic incidence rate itself. We calculated cross-correlations between the traffic timelines and the epidemic timelines, to understand in which states traffic had a tighter coupling with the epidemic. Since one would expect that existing properties of the state (such as density, political affiliation) are factors contributing to the strength of this coupling, we investigated whether the degree of coupling correlated with any of the state demographic measures.

A subsequent direction of study was to ask to what extent high coupling between the epidemic and traffic timelines reflected into poor epidemic outcome for the corresponding states; in other words, to what extent high mobility during high infection periods contributed to refueling the epidemic, and which were the traffic modalities most associated with the epidemic increase.

Finally, we investigated the potential that traffic increased in regions where vaccines were prevalent, possibly as an effect of people gaining confidence in the added protection confered by the vaccine. To do this, we calculated correlations between the efficiency of vaccinations and the degree of traffic since January 2020.

## 3 Results

### 3.1 State demographic profiles and epidemic evolution

We computed Spearman correlations between %*CI* and all measures in the demographic profiles of the states. The significant correlation values are reported in Tables 1 and 2.

**Table 1:** Correlations between epidemic measures and state demographic profiles as reflected in population density, median household income (MHI) and political affiliation (% blue votes). For significant correlations, both the Spearman correlation value and the significance p-value are shown, in separate columns.

|  | Density |  | MHI |  | % blue votes |  |
| --- | --- | --- | --- | --- | --- | --- |
|  | Corr coef | p-value | Corr coef | p-value | Corr coef | p-value |
| %CI total | – | – | -0.28 | 0.04 | -0.47 | $< 10^{-3}$ |
| %CI wave 1 | 0.51 | $< 10^{-3}$ | 0.27 | 0.05 | 0.40 | $< 10^{-2}$ |
| %CI wave 2 | – | – | -0.47 | $< 10^{-3}$ | -0.48 | $< 10^{-3}$ |
| %CI wave 3 | -0.51 | $< 10^{-3}$ | -0.30 | 0.03 | -0.61 | $< 10^{-5}$ |
| %CI wave 4 | 0.46 | $< 10^{-3}$ | – | – | – | – |
| %CI wave 5 | 0.38 | $< 10^{-2}$ | 0.30 | 0.02 | 0.36 | $< 10^{-2}$ |

**Table 2:**
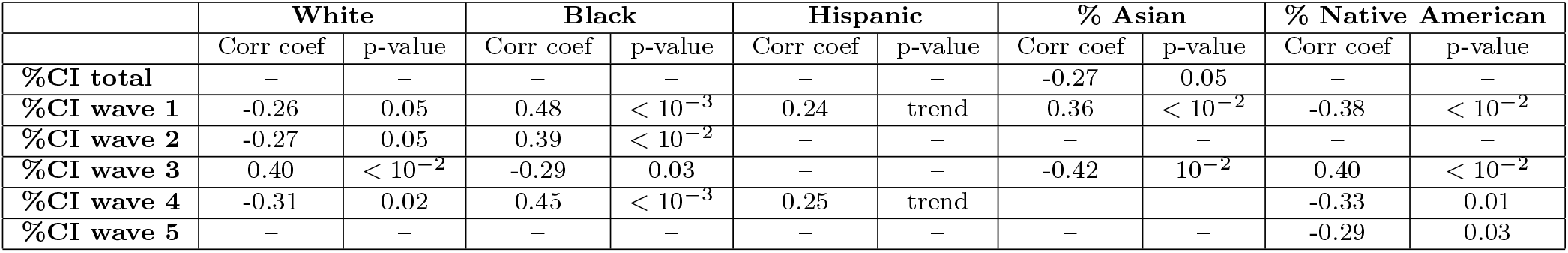
Correlations between epidemic measures and the state’s racial/ethnic profile. For significant correlations, both the Spearman correlation value and the significance p-value are shown, in separate columns.

Since we hypothesized that different epidemic waves (as defined in the Methods section) may have had different driving mechanisms, we also calculated the same correlations, using the %*CI* computed separately for each wave (with the significant results reported in Tables 1 and 2). Notice that each epidemic wave exhibits a different signature of correlations with the state profiles, and that these signatures are very different between waves. For example, the values in Table 1 show that, for the first wave, the magnitude of its epidemic aftermath (measured as the wave’s %*CI*) was positively correlated with population density, MHI and percent of blue votes (in other words, denser, wealthier, blue states were more affected). The third wave had the exact opposite signature, with the %*CI* negatively correlated with population density, MHI and percent of blue votes (in other words, denser, wealthier, blue states were less affected).

Such differences persist in the correlations between the waves’ epidemic outcome and the racial/ethnic profiles of the corresponding states. Here again, the first and third wave show opposite trends: the first wave was significantly less pronounced in states with a higher white population, and more pronounced in those with higher black population (as shown by the negative and respectively positive correlation values in the corresponding row of Table 2). In contrast, the third wave was significantly more pronounced in states with a higher white population, and less pronounced in those with higher black population (as shown by the positive and respectively negative correlation values in Table 2).

Notice that some of these effect cancel out to a large degree when considered over all waves together, making it seem that there is no correlation between the total %CI and the state profile measures, when in fact different (often opposite) correlations emerged for different portions of the epidemic timeline. These correlations may be the sign of different mechanisms governing each epidemic wave, and will be further discussed in our last section.

### 3.2 State demographic profiles and vaccination timelines

We showed in the Methods section that vaccination levels differed widely between states. A natural question to ask is to what extent vaccination efficiency of a state depended on (and could have been predicted from) its demographic profile. This may be very important information when aiming to optimize the impact and outcome of vaccination for the incoming waves of the current epidemic, or for vaccination campaigns in general.

As a first step, we computed correlations between the percent of fully vaccinated people in each state by May 22, 2021, and all demographic measures in our profile. We found strong positive correlations with the state’s density, median household income and percent of blue votes in the 2020 presidential election (as shown in Table 3). This is not a surprise, but may not be trivial to interpret. For example, high density may be in and of itself an incentive to vaccinate (since it brings higher exposure rate), but may also be associated with high urbanization, politics, MHI and other demographic aspects, through which it may indirectly affect vaccination rates. Similarly, income may be related with education level, and thus may indirectly affect people’s decisions with respect to whether or not to vaccinate. Finally, politics is likely to influence vaccination schedules directly via state government measures and policies (which varied widely between blue and rd states). All these ideas will be further explored in the Discussion.

**Table 3:** Correlations between vaccination measures and state demographic profiles as reflected in population density, median household income (MHI) and political affiliation (% blue votes). For significant correlations, both the Spearman correlation value and the significance p-value are shown, in separate columns.

|  | Density |  | MHI |  | % blue votes |  |
| --- | --- | --- | --- | --- | --- | --- |
|  | Corr coef | p-value | Corr coef | p-value | Corr coef | p-value |
| %CV | 0.30 | 0.03 | 0.63 | $< 10^{-5}$ | 0.76 | $< 10^{-10}$ |
| % drop in %DV since peak | -0.53 | $< 10^{-4}$ | -0.63 | $< 10^{-5}$ | -0.73 | $< 10^{-10}$ |
| % drop in %DV since April 13 | -0.51 | $< 10^{-3}$ | -0.63 | $< 10^{-6}$ | -0.73 | $< 10^{-8}$ |

Our second correlation analysis focused on the relationship between the states’ profiles and the size of the drop in vaccination rates after the Johnson & Johnson pause on April 13. The percent change in %*DV* (measured either from the peak rate for each state, or from April 13) showed a significant inverse correlation with the state’s density, median household income and percent of blue votes in the 2020 presidential election (also shown in Table 3). This is an interesting effect, and supports the idea that the vaccination rate drop after the J&J incident is not accidental, but rather a behavioral response, modulated by each state’s individual profile. Again, political affiliation of the state seems to be one of the driving factors in shaping this response, and may be an effect to consider for future measures of epidemic mitigation.

In order to observe vaccination patterns for different racial/ethnic profiles (at the state level), we computed the correlations of the same vaccination measures with the % of the state’s population in each of the five ethnic groups described in the Methods section. The results are shown in Table 4. Our strongest correlations came across for states’ association with higher Asian population: in these states, the cumulative full vaccinations were higher, and the percent drop after April 13 was less dramatic, suggesting a more efficient response to vaccinations policies. The drop was also significantly lower in states with higher Hispanic population, but significantly higher in those with higher Native American population. Cumulative vaccinations were found to be marginally lower in states with higher Black population. These results will be further explained and contextualized in our Discussion section.

**Table 4:**
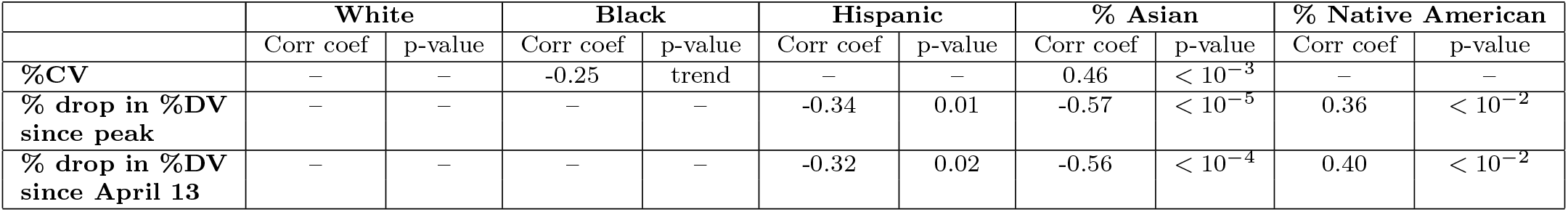
Correlations between vaccination measures and the state’s racial/ethnic profile. For significant correlations, both the Spearman correlation value and the significance p-value are shown, in separate columns.

|  | White |  | Black |  | Hispanic |  | % Asian |  | % Native American |  |
| --- | --- | --- | --- | --- | --- | --- | --- | --- | --- | --- |
|  | Corr coef | p-value | Corr coef | p-value | Corr coef | p-value | Corr coef | p-value | Corr coef | p-value |
| %CV | – | – | -0.25 | trend | – | – | 0.46 | $< 10^{-3}$ | – | – |
| % drop in %DV since peak | – | – | – | – | -0.34 | 0.01 | -0.57 | $< 10^{-5}$ | 0.36 | $< 10^{-2}$ |
| % drop in %DV since April 13 | – | – | – | – | -0.32 | 0.02 | -0.56 | $< 10^{-4}$ | 0.40 | $< 10^{-2}$ |

A last natural question to ask is whether vaccinations appear to have efficiently contributed to epidemic control by May 22, 2021 (when relating the two at the state level). As a first step in this direction, we computed correlations between epidemic control and vaccination efficiency, between January 8 (the first day in our state-wide vaccination data set) and May 22, 2021. As a measure of epidemic control, we used the percent change in cumulative incidence; as a measure of vaccination efficiency, we used the total number of vaccines administered during the period of interest. Since vaccines do not reach full protection until 2 weeks after completion of full vaccination (in average), we considered a 2 week lag between the window for our vaccination measure (January 8 until May 8, 2021) and the window for our epidemic measure (December 25, 2020 until May 22, 2021). The two measures were weakly correlated, with a Spearman correlation value of *ρ* = 0.30, and a significance of *p* = 0.02.

### 3.3 State demographic profiles and traffic timelines

Our first objective was to establish whether there are correlations (1) between the average level of traffic in each state (as percent of baseline) and any of the state’s demographic measures, on one hand, and (2) between the traffic level and the epidemic outcome in the state, on the other.

For (1), we computed the average traffic levels for all three modalities (driving, public transit and walking) between January 13, 2020 and May 22, 2021 (with respect to each state’s baseline). and computed correlations with all measures in the states’ demographic profile. We obtained weak correlations, as follows. The public transit showed a negative correlation with the MHI (*ρ* = −0.42, *p* = 0.004) and with the percent of blue votes (*ρ* = −0.33, *p* = 0.02); it was also negatively correlated with the percent of Hispanic population (*ρ* = −0.32, *p* = 0.03) and of Asian population (*ρ* = −0.42, *p* = 0.003). We found average walking to be negatively correlated with the state’s population density (*ρ* = −0.34, *p* = 0.01) and positively correlated with the percent of Native American population (*ρ* = 0.53, *p* < 10^−4^). No correlations with state profile measures were found for average driving.

Some of these results suggest common sense potential explanations. For example, the inverse correlation between the level of public transit and MHI could reflect the fact that in wealthier states jobs allowing remote work may have been more prevalent, or people may have been able to avoid public transit by other means (analysis which is not included in this study). Blue political affiliation may associate with lower transit via stricter social distancing mandates, although other, indirect (assessment-related) factors may contribute (such as the fact that blue states may have more urban areas, and more developed public transit, hence a higher baseline to begin with). It is also not surprising that the walking average correlated negatively with the states’ density, since sparser population can be seen as an incentive for people to feel safer walking without exposure.

For (2), we considered, as before, the average traffic levels (as baseline percentages) for all three modalities between January 13, 2020 and May 22, 2021, and we computed %*CI* for the same time period. We obtained no significant correlations between traffic end epidemic measures, for any of the three modalities. However, this does not necessarily mean that traffic had no contribution to the epidemic dynamics. These comparisons are very coarse, since they use one bulk measure (average traffic) to describe a time series in which fluctuations, and not just the mean, hold very high relevance. In particular, one important question is how the *fluctuations* in traffic synchronized with those in epidemics. In general, one would expect high mobility close to the peak of an epidemic wave to have a more detrimental epidemiological impact than high mobility when infection is low. A lot of recent discussions have been around the fact that different states had very different approaches to this synchronization.

To approach this idea rigorously, we first quantified the synchronization of traffic and epidemic timelines by computing cross-correlations *CCR* of traffic and percent daily incidence timelines (as illustrated in Figure 6a). As expected, states had many different approaches to limiting traffic during periods of increased infection rates, resulting into a wide distribution of *CCR* values, as illustrated in the histogram in Figure 6b for driving (for all 50 states for which Apple provided driving information). Interestingly, the figure shows that the highest *CCR* value was achieved by North Dakota, which (as we previously shown) also had the highest %*CI*. We are interested to understand whether this is coincidental, or if it is representative of general statistics of high *CCR* being associated with poor epidemic outcome (high %*CI*). If this is true, we are also interested to understand what are the state-wide factors that may have lead to people traveling during periods of high infection, hence high *CCR*. To these aims, we rephrase our two questions to include this timing component, by looking for correlations (1) between the values of *CCR* for the states, and their epidemic outcome, on one hand, and (2) between *CCR* and the states’ demographic measures, on the other.

**Figure 6:**
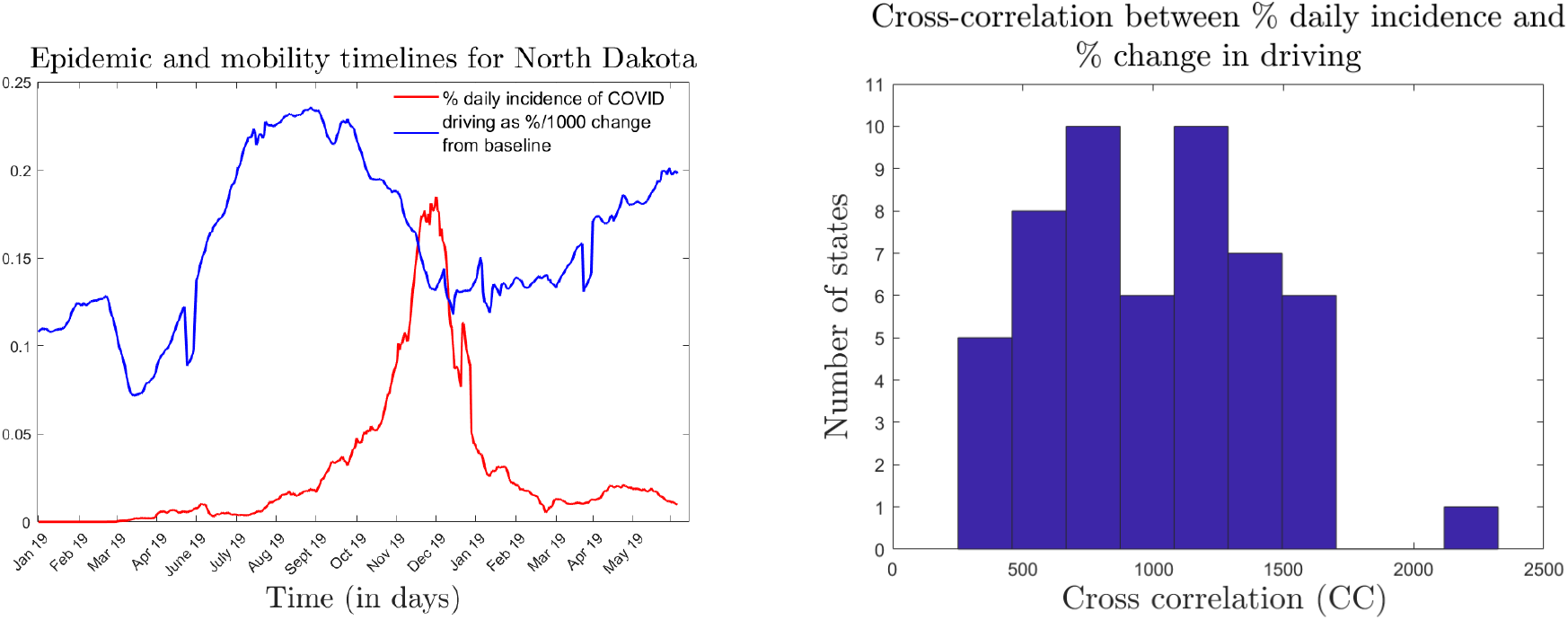
Cross-correlations (CCR) between driving and epidemic timelines. **A**. *Comparison between the fluctuations in epidemics (measured as* %*DI) and in driving levels (visualized as 0*.*001% baseline, to facilitate visual comparison) for the state of North Dakota. The cross-correlation (CCR) of the two time series is shown on the right*. **B**. *Histogram illustrating the distribution of the CCR values (between driving levels and* %*DI) for all 50 states for which driving information was available. The high CCR outlier represents the state of North Dakota*.

We found that high *CCR* for all three traffic modalities correlated positively with poor epidemic outcome: *ρ* = 0.51 and *p* < 10^−3^ (for the 50 states for which driving information was available); *ρ* = 0.41 and *p* = 0.005 (for the 43 states for which public transit information was available); *ρ* = 0.61 and *p* < 10^−5^ (for the 50 states for which driving information was available). Scatter plots for all three correlation statistics are shown in Figure 7, for illustration.

**Figure 7:**
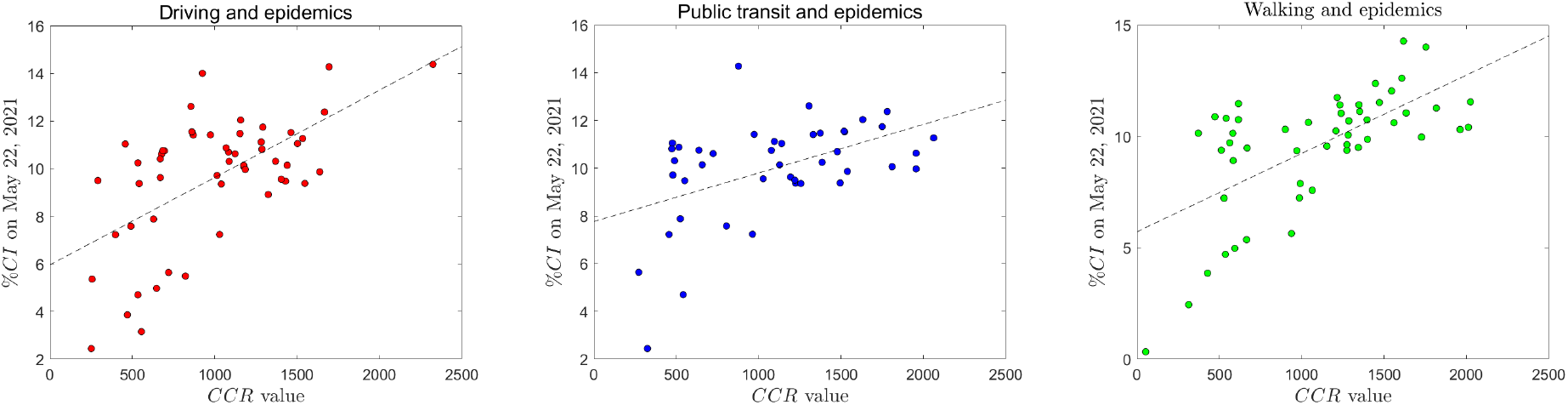
Scatter plots. *illustrating correlations between CCR values for all three traffic modalities (***A**. *driving;* **B**. *public transit;* **C**. *walking), with the states’ epidemic outcome (measured as* %*CI)*.

This reinforces the idea that states which allowed high population mobility when infection rates were high, hence the risk of exposure was increased, were more likely to end up with an elevated percent cumulative incidence. However, the question of causality remains, and cannot be simply answered by a correlation analysis. Instead, we aimed to establish next the potential bases for mobility patterns leading to high *CCR* values, by investigating whether the latter related statistically to any of the states’ demographic measures. The results of the correlation analysis between *CCR* values and demographic measures are shown in Table 5. Notice that the correlations have different signs depending on the traffic modality. Potential underlying explanations will be discussed in our last section.

**Table 5:** Correlations between vaccination measures and state demographic profiles as reflected in population density, median household income (MHI) and political affiliation (% blue votes). For significant correlations, both the Spearman correlation value and the significance p-value are shown, in separate columns.

|  | Density |  | MHI |  | % blue votes |  |
| --- | --- | --- | --- | --- | --- | --- |
|  | Corr coef | p-value | Corr coef | p-value | Corr coef | p-value |
| **Driving CCR** | 0.30 | 0.03 | 0.63 | $< 10^{-5}$ | 0.76 | $< 10^{-10}$ |
| **Transit CCR** | -0.53 | $< 10^{-4}$ | -0.63 | $< 10^{-5}$ | -0.73 | $< 10^{-10}$ |
| **Walking CCR** | -0.51 | $< 10^{-3}$ | -0.63 | $< 10^{-6}$ | -0.73 | $< 10^{-8}$ |

## 4 Discussion

In our paper, we proposed and analyzed different factors that may have contributed to the evolution of the COVID-19 outbreak in the US, since its start in January 2020, until May 2021. We considered time-dependent factors tightly coupled with COVID-19 incidence (such as population mobility and vaccinations), but also underlying factors which remained unchanged for the duration of the pandemic (such as density, income, political affiliation by state). Association with the latter can help generate epidemic predictions based on existing state profiles. The former may help explain the mechanisms underlying predictable patterns in epidemic trends.

First of all, our results reiterate a previously explored idea that epidemic, vaccination and traffic trends varied widely across the US states. Our correlation analyses show that some of these differences may stem from demographic factors. For example: while COVID-19 daily incidence timelines showed overall five different epidemic waves, the subset of waves experienced by each state, and their amplitudes, varied widely across states. With our analysis, we first showed that each state’s overall COVID-19 incidence was lower in states with higher income and stronger blue political affiliation, and did not appear to be correlated with the state’s population density, or with its ethnic/racial profile. However, we also found that this did not come across as a consistent trend along the epidemic timeline, and that different waves had completely different, strong correlations with some of these factors.

For example, we pointed out that the first wave affected more (in terms of COVID-19 incidence) denser, wealthier, bluer states; it also disproportionately affected states with higher Black, Hispanic and Asian populations, while states with higher White populations were less affected. A possible explanation for these relationships is that the first wave was notoriously characterized by unpreparedness. Higher income was not immediately relevant in the middle of a resource crisis (it took time to obtain tests and PPE, even when afforded by the state), and neither was political affiliation. Rather, the epidemic spread fast in states with higher density, more urban, with more travel hubs (the states with earliest infection being New York, California, Washington). While these were primarily blue states, political affiliation only became relevant at the point where mitigation strategies were clear, and their implementation started to depend on governmental measures. Before such measures were introduced, took hold and eventually started to show effects, it is likely that the wave was driven primarily by the intrinsic characteristics of the state that directly impact exposure, (such as density, connectivity, geographic location), with an uneven distribution of the very limited resources among ethnic/racial groups.

These patterns started to swap over the summer (second) wave, and eventually settled to the exact opposite correlations for the fall wave, when infections was more rampant in states with lower density, income and blue affiliation. The negative correlation with MHI suggests that wealthier states may have better afforded mitigation strategies; however, it is interesting to note in this context that states with a higher percent of White population were more affected, and those with higher Black population were less affected. This suggests that the behavioral population response (rather than resources) may have been a stronger determinant of the spread in this epidemic wave. One possible explanation is that these states had less political incentive to abide by rules minimizing exposure (there have been many discussions around the fact that state-governmental mandated lockdowns and social distancing strategies were more efficiently implemented in states with blue government [21]).

One way to further investigate this idea was to ask how the population mobility can be associated with exposure risk, and whether this depended on the political color of the state. For each state, we compared traffic levels (in driving, public transit and walking, as made available by Apple) with the state’s cumulative incidence by May 22, 2021. No significant correlations were obtained between these two overall measures, for any of the three modalities. We refined the analysis to include the relevant *timing* component, which accounted for the important aspect of people traveling during periods of high versus low infection in the state. To capture this aspect, we computed lag zero cross-correlations between the traffic and the epidemic time courses between January 2020 and May 2021; we interpreted higher cross-correlation values as higher level of traffic during high infection windows. We then showed that higher cross-correlations were significantly associated with poorer epidemic outcomes (higher cumulative incidence) for all three traffic modalities, supporting the idea that the timing of travel may have been crucial to the spread of COVID-19 (irrespective of the travel modality).

Correlations with the state’s demographic profile (including political affiliation) differed by modality, with higher driving-epidemic coupling, but lower transit-epidemic and walking-epidemic coupling in denser, wealthier and bluer states. Out of all three modalities, a potential direct interpretation can only be easily generated in the case of transit, where the transportation itself may represent (and may be perceived as) a source of exposure to the SARS-Cov2 virus. In that case, as expected, the level of transit showed increased synchronization with the epidemic waves in states with lower density, lower MHI, and lower percent of blue votes. Interpretation of the correlations corresponding to driving and walking is more complicated. While driving and walking don’t represent high exposure behaviors in and of themselves, the data contain no knowledge of *where* individuals traveled to (e.g., high risk versus low risk destinations), and under what circumstances (car pooled with others, etc). A possible explanation of the positive correlations in the case of driving is that in denser, wealthier, bluer states people may have been additionally motivated (and able) to switch more transportation methods to driving when infection increased, resulting in higher *CCR* values (which did not necessarily mean higher contamination risk). However, this explanation does not agree with our previous result, showing that higher driving *CCR* values were in fact correlated with poorer epidemic outcome, suggesting a potentially significant epidemiological contribution of driving during high infection waves.

Finally, we investigated the evolution and potential role of vaccinations to the dynamics of the epidemic system. We found that, beyond differences between states, all daily vaccination timelines showed a highly correlated shape peaking around April 12 – the day the administration of the Johnson and Johnson vaccine was paused by the CDC and FDA, based on reported clinical side-effects. While administration of J&J vaccines was resumed on April 23, daily vaccination rates continued to drop until May 22 (the last day of our study), suggesting that this drop did not simply reflect the temporary absence of J&J vaccinations, or people switching to other vaccines, but rather a longer-term behavioral response of the population to the perceived risk of vaccine side-effects. Interestingly, this trend seems to have only affected US states (and not other countries, which had experienced similar issues, e.g. with the Astra Zeneca vaccine, in most European countries).

Despite the same up-down shape in vaccination daily rates, states overall efficiency varied widely, between 26-51% by May 22. We found that, by May 22, the percent of fully vaccinated people in each state’s correlated strongly with density, HMI and political affiliation, suggesting that people’s decision to vaccinate may have depended on their assessment of exposure risk (density), on the state’s income status (related to either vaccine availability, or possibly education status, not analyzed in this study), and on the state’s politics (via governmental incentives to vaccinate, but also possible through other correlates). In addition, the same factors (and likely the same explanations) related to the degree of of the drop in vaccination rates after the J&J incident (as computed from April 12 until May 22), which varied between 10% and 83%, with lesser drops in denser, wealthier, bluer states. It in notable that the level of cumulative vaccinations by May 22 did correlate with the percent change in cumulative incidence of COVID-19 (a weak, although significant correlation, which might improve if more time were allowed).

Altogether, our study supports the idea that the significant differences in epidemic, vaccination and mobility measures across the US states may have at least partly stemmed from demographic factors, and from political differences. In particular, the population’s behavioral response to the epidemic fluctuations influenced mobility and vaccinations differently across states. Some of these differences can also be explained by differences in demographic measures and political aspects (reflected in state government mandates). This is very important, because these differences in turn likely influenced the state-wide epidemic outcome.

## 5 Limitations and future work

Our study has some limitations intrinsically related to epidemic data collection and reporting. In terms of epidemic data, there have been many discussions around artifacts in daily reporting of COVID-19 cases. Some of these (such as weekly fluctuations) can be eliminated via pre-processing, others are more difficult to locate and resolve. In terms of traffic data, a substantial limitation comes from the target destination not being identified in the Apple data, hindering the process of associating clear exposure risk to traffic information. This could be improved in future work by including additional, destination-specific data (as provided by Google). The data on vaccines lacks follow-up assessment of the efficacy and conferred immunity timeline (which cannot yet be evaluated clinically), laying questions on the efforts of estimating epidemic impact at different time points. Considering time series past May 22, as well as a better understanding of vaccine-induced immunity would help refine this analysis.

Other limitations raise from our study design, which uses a limited subset of (possibly not independent) measures, reports state-wide results, and uses simple correlation analyses. While this setup is valuable as a first step, and establishes a proof of principle, further work should focus on expanding the set of demographic measures (to include, for example, geographic location, education level, urbanization). State-level results can be easily refined at the county level (as done in one of our previous studies on New York State alone), adding to the spatial coarse graining, but also to the statistical power. Finally, the current correlation analysis can be used to guide a more detailed factor analysis, which would improve the approach to selecting the relevant factors.

## Data Availability

All data was accessed from the public domain, and all sources are referenced in the manuscript.

## 6 Acknowledgments

We would like to thank Dr. Nancy Campos for the continuing support for carrying our this project.

## Notes

### Competing Interest Statement

The authors have declared no competing interest.

### Funding Statement

This study was supprted by the SUNY New Paltz AC2 summer program.

